# A deep convolutional neural network-based algorithm for muscle biopsy diagnosis outperforms human specialists

**DOI:** 10.1101/2020.12.15.20248231

**Authors:** Yoshinori Kabeya, Mariko Okubo, Sho Yonezawa, Hiroki Nakano, Michio Inoue, Masashi Ogasawara, Yoshihiko Saito, Jantima Tanboon, Luh Ari Indrawati, Theerawat Kumutpongpanich, Yen-Lin Chen, Wakako Yoshioka, Shinichiro Hayashi, Toshiya Iwamori, Yusuke Takeuchi, Reitaro Tokumasu, Atsushi Takano, Fumihiko Matsuda, Ichizo Nishino

## Abstract

Histopathologic evaluation is essential for categorizing and studying neuromuscular disorders. However, experienced specialists and pathologists are limited, especially in underserved areas. Although new technologies, such as artificial intelligence, are expected to improve medical reach, their use in rare diseases is challenging because of the limited availability of training datasets. To address this knowledge gap, we developed an algorithm based on deep convolutional neural networks that used data from microscopic images of hematoxylin-and-eosin-stained pathology slides. Our algorithm differentiated idiopathic inflammatory myopathies (mostly treatable) from hereditary muscle diseases (mostly non-treatable) and achieved better sensitivity and specificity than real physicians’ diagnoses. Furthermore, it successfully and accurately classified four subtypes of the abovementioned muscular conditions. These results suggest that our algorithm can be safely used in a clinical setting. We established the similarity between the algorithm’s and physicians’ predictions using visualization technology, and clarified the validity of the predictions.

## MAIN

### Introductory Paragraph

The diagnosis, management, and further study of rare diseases carry fundamental challenges that are different from those of common diseases, owing to fewer patients and limited expert facilities and clinicians^1^. Novel digital tools, such as artificial intelligence (AI), are expected to circumvent these shortcomings by accelerating the processes of diagnosis, specialist referrals, gathering and sharing of data, and clinical research on rare diseases^2^. Deep learning is a highly reliable AI technology used for specific tasks^3^; analyzing medical and pathological images using deep learning^4-6^ is comparable to that by human experts^6-9^. However, almost all deep learning-based medical image analyses have, so far, only dealt with common diseases^6-9^ due to the limited data available on rare diseases. Previous studies using muscle MRI and AI scored the conditions manually and did not diagnose them using the images directly^10,11^.

In this study, we aimed to histopathologically diagnose rare muscle diseases. Evaluating muscle pathology is unique because muscle biopsy specimens require freeze-fixation and a different set of histochemical stainings. Classifying muscle diseases according to their pathological features remains diagnostically relevant even in today’s molecular-diagnosis era^12^.

Previous studies required whole-slide pathological images^13,14^ and significant infrastructural investments. However, real-world pathological diagnoses are performed with analog microscopes. Developing a diagnostic tool with a CCD camera that is much cheaper than a whole slide scanner and can recruit analog microscope images is the key to establishing a practically applicable system, especially in underserved areas worldwide.

To address this issue, we developed a deep learning, convolutional neural networks (CNNs)-based algorithm that could differentiate between major muscle diseases using a small amount of training data. To train and evaluate the algorithm, we collected microscopic images of hematoxylin and eosin (H&E)-stained pathological slides that were obtained by charge-coupled device (CCD) cameras. The underlying algorithmic architecture for classifying dominant muscular dystrophies with whole-slide images has been established^15^.

We chose 11 muscular diseases; 4 idiopathic inflammatory myopathies (IIM) [dermatomyositis (DM), inclusion body myositis (IBM), immune-mediated necrotizing myopathy (IMNM), and antisynthetase syndrome (ASS)] and 7 hereditary muscle diseases, [dystrophinopathy (DYST), limb-girdle muscular dystrophy 2A (LG2A), limb-girdle muscular dystrophy 2B (LG2B), Ullrich congenital muscular dystrophy (UCMD), Fukuyama-type congenital muscular dystrophy (FCMD), congenital myopathy (CM), and GNE myopathy (GNEM)] (**Table 1**).

**Table 1.** Summary of studies, diseases, and the number of pathological images and slides. The number of images obtained from one slide was mostly 3, but it could vary (1– 6) depending on the size of tissue on the slide.

| Group | Disease | Images/Slides |
| --- | --- | --- |
| Idiopathic<br>inflammatory<br>myopathy<br>(IIM) | DM | 307/103 |
|  | IBM | 306/102 |
|  | IMNM | 332/109 |
|  | ASS | 317/107 |
| Non-myositis<br>muscle disease | DYST | 348/126 |
|  | FCMD | 256/95 |
|  | LG2A | 321/120 |
|  | UCMD | 213/83 |
|  | LG2B | 512/177 |
|  | CM | 629/211 |
|  | GNEM | 181/60 |
| Others | NP | 319/107 |
| Total |  | 4041/1400 |

We employed a two-step approach to distinguish between the diseases: 1) differentiating IIM from other conditions, and 2) subclassifying each category because most IIM conditions are treatable and other hereditary conditions are not (**Figure 1a)**. In the first step, we combined images of DM, IBM, IMNM, and ASS to create the IIM group, and those of DYST, FCMD, LG2A, LG2B, UCMD, CM, GNEM, and NP to create the counterpart group. We used a holdout method^16^ to train and evaluate CNNs and compare them with human physicians. In the second step, we subclassified 4 IIM subtypes and 7 hereditary muscle diseases. We evaluated the results using 5-fold cross-validation.^16^

**Figure 1.**
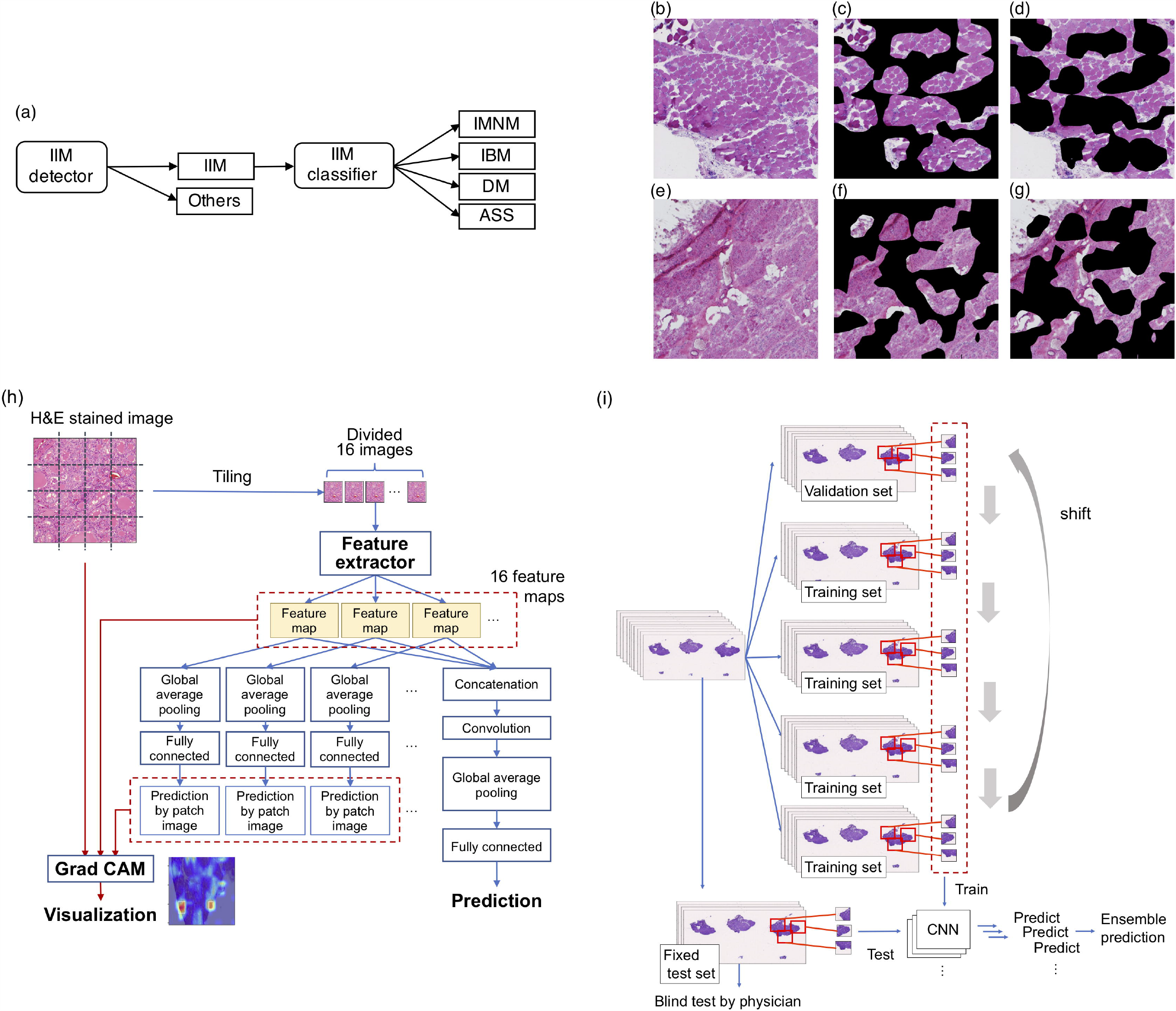
Strategy, masked sample images, and deep convolutional neural network architecture. (a) The strategy of IIM identification, which consists of differentiating IIM from other diseases and classifying the subtypes of IIM. (b) An IIM (IMNM) image. (c) AI-focused image with masked areas not to be focused by CNNs in (b). (d) AI-unfocused image with masked areas to be focused by CNNs in (b). (e) A non-myositis muscle disease (FCMD). (f) AI-focused image with masked areas not to be focused by CNNs in (e). (g) AI-unfocused image with masked areas to be focused by CNNs in (e). (h) Deep CNN architecture; blue arrows indicate the flow for training and prediction, and red arrows indicate the visualization flow. (i) Approach for training and evaluating CNNs as compared with those of the physicians. The images were divided beforehand into a training set and a test set. The training set was divided into five groups; four were used for training, and one was used to check the progress of the training as a validation set. The training was executed five times while shifting the validation set one by one, and all five CNNs were trained.

We also developed a visualization method using Grad-cam^17^ to check the CNNs’ prediction accuracy. We used it to create AI-focused images (**Figure 1c and 1f**) that masked the areas that were not evaluated by the CNNs and AI-unfocused images (**Figure 1d and 1g**) that masked the areas to be targeted by CNNs. We then identified the image group that physicians could use to obtain correct diagnoses and investigated the relationship between the CNNs’ predictions and the physicians’ diagnoses.

The CNN architecture developed for this study is illustrated in **Figure 1h**. The CNNs divided an imported image into 16 patches and generated a feature map per patch. The 16 feature maps were concatenated and used for the prediction of each image, due to which we expected that the CNN could be trained effectively even with a small number of images. The CNN calculated the probability for each disease, and the highest probability was considered the predicted class. Further, we used a ‘model ensemble’ method^18^ to improve the CNNs’ performance in differentiating IIM from other conditions (**Figure 1i**).

The deep CNN was able to precisely differentiate IIM from other muscle diseases with an area under the curve (AUC) of 0.996 and outperform 9 human specialists (**Figure 2a**). The average and the inter-variability of 9 human specialists’ accuracies were 83.4% and 0.004, respectively. Its accuracy was 96.9% if a probability of 0.5 or higher was determined as myositis, while the physicians’ highest accuracy was 93.8%. The average and the variability of 9 human specialists’ accuracies were 83.4% and 0.004, respectively. The number of misclassifications was similar between IIM and other conditions (**Figure 2b**). If a probability of 0.3 or more was determined as IIM, the accuracy decreased to 95.8% but the number of incorrectly classified IIMs became 0 (**Figure 2c**). We also confirmed that the ‘model ensemble’ was efficient in further improving the CNNs’ performance (**Figure 2d**).

**Figure 2.**
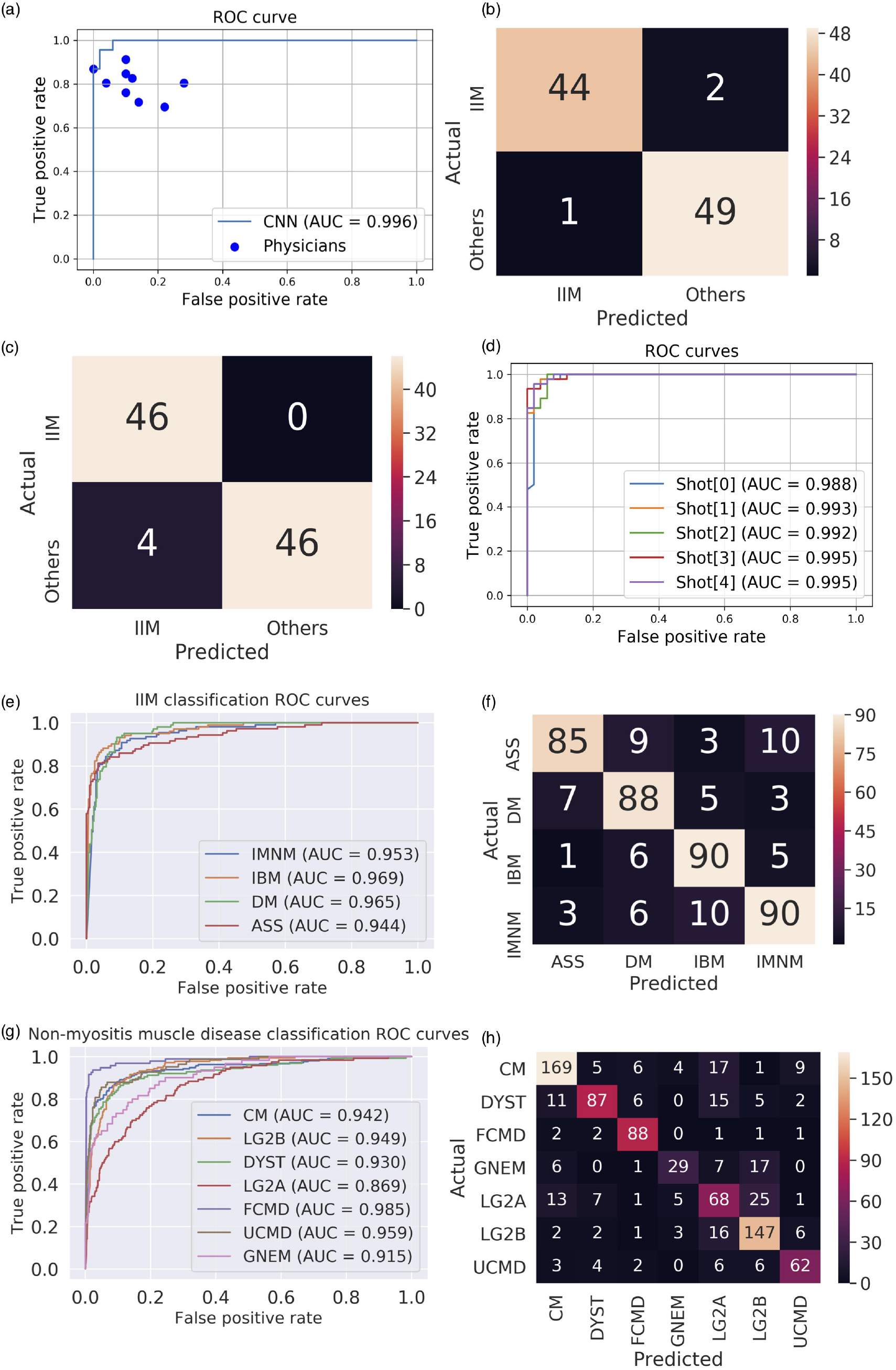
Differentiation of IIM and classification of IIM and non-myositis muscle disease. (a) A ROC curve of the CNN (blue line) and physicians’ (blue dots) performances. (b) The confusion matrix that separates the IIM from the other diseases is 0.5. The vertical is the actual state and the horizontal is the prediction. (c) The confusion matrix that separates the IIM from the other diseases is 0.3 and there were no misclassifications of IIM. (d) ROC curves without ensemble methods. (e) ROC curves of IIM classification; the blue line shows IMNM, the orange line is IBM, the green line is DM, and the red line is ASS. (f) Confusion matrix of IIM classification. The vertical is the actual state and the horizontal is the prediction. (g) ROC curves showing the classification of non-myositis muscle disease. The blue line indicates CM, the orange line is LG2B, the green line is DYST, the red line is LG2A, the purple line is FCMD, the brown line is UCMD, and the pink line is GNEM. (h) Confusion matrix of non-myositis muscle disease classification.

The CNN successfully classified 4 subtypes of IIM with AUCs being 0.953 (IMNM), 0.969 (IBM), 0.965 (DM), and 0.944 (ASS) (average = 0.958) (**Figure 2e**). We found that the accuracy was 83.9% when the disease with the highest probability was the predicted class; ASS tended to be incorrectly classified as IMNM; and IMNM tended to be erroneously judged as IBM (**Figure 2f**). It also classified 7 subtypes of hereditary muscle disease with AUCs of 0.942 (CM), 0.949 (LG2B), 0.930 (DYST), 0.869 (LG2A), 0.985 (FCMD), 0.959 (UCMD), and 0.915 (GNEM) (average = 0.936) (**Figure 2g**). We found that the accuracy was 74.5% when the disease with the highest probability was the predicted class; LG2A tended to be classified as LG2B; and GNEM tended to be misjudged as all other classes (**Figure 2h**).

Red and yellow areas were the most important to visualize the CNN predictions (**Figure 3b, 3d**) because they were interspersed within the specimen (**Figure 3a-3d**). The average accuracy of the physicians’ diagnosis with AI-focused and AI-unfocused images in myositis was 0.674 and 0.526, respectively (**Figure 3e**). The accuracy of DM decreased relatively significantly (**Figure 3f and 3g**). The average accuracies of non-myositis diseases were 0.458 and 0.440, respectively (**Figure 3h**). Comparatively, the AI-focused and AI-unfocused images of myositis conditions were significantly different (p = 0.003) but those of non-myositis conditions were not (p = 0.629) (**Figure 3f-g and 3i-j**).

**Figure 3.**
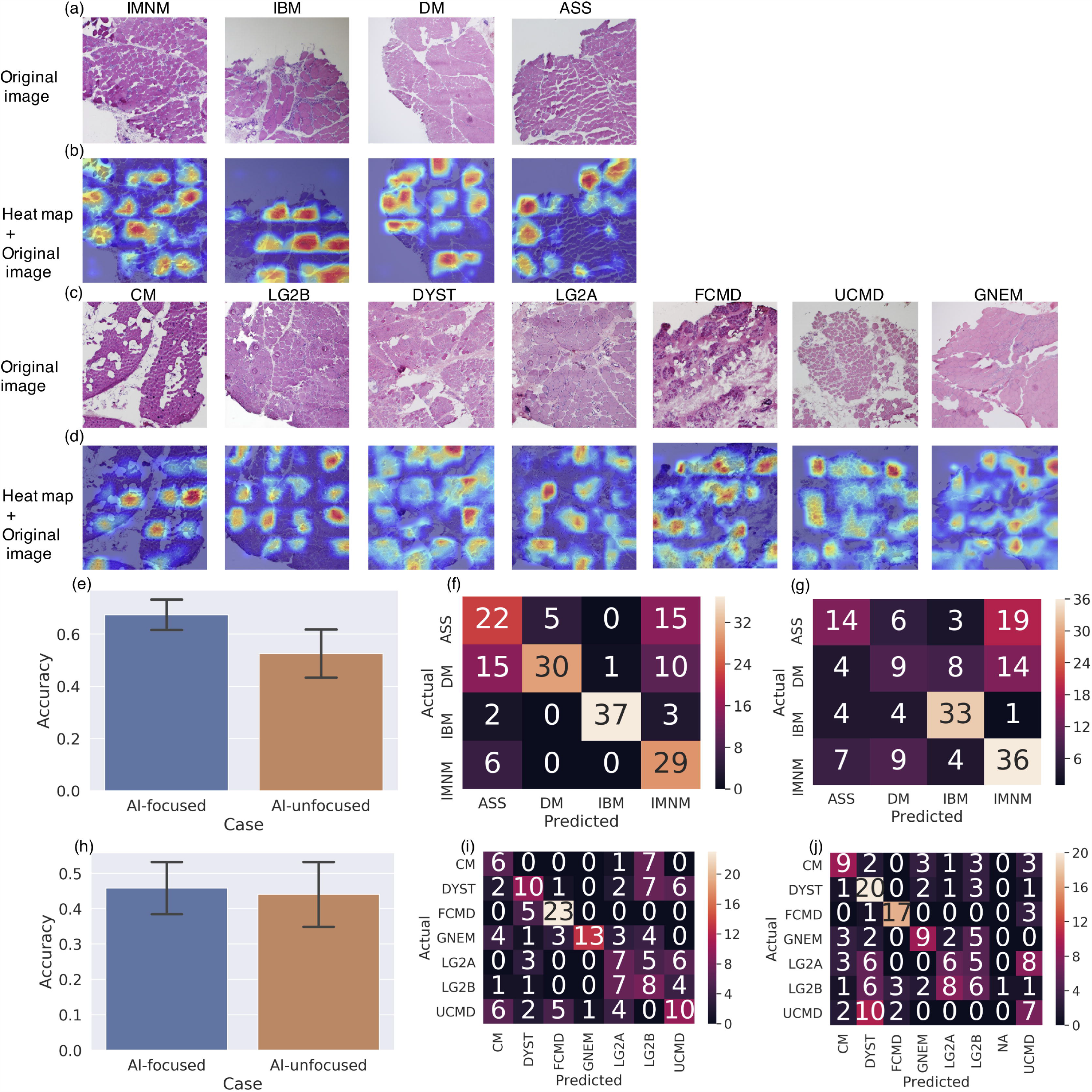
Visualization of CNNs predictions. (a) Sample H&E-stained images of IIM. Left to right: IMNM, IBM, DM, and ASS. (b) Merged images of H&E-stained images and heatmap images created with Grad-cam. Red color indicates CNNs-focused areas as they are essential for the prediction. (c) Sample H&E-stained images of non-myositis muscle diseases. Left to right: CM, LG2B, DYST, LG2A, FCMD, UCMD, and GNEM. (d) Merged images of H&E-stained and heatmap images created with Grad-cam. (e) Physicians’ test results in myositis. The left (blue) bar shows the average accuracy of the physicians’ diagnosis with AI-focused images, while the right (orange) bar shows the AI-unfocused images. The black line indicates the standard deviation. (f) Confusion matrix of results with AI-focused images in IIM. (g) Confusion matrix of results with AI-unfocused images in IIM. (h) Physicians’ test results in non-myositis. The left (blue) bar shows the average accuracy of the pathologists’ diagnosis with AI-focused images, and the right (orange) bar shows the AI-unfocused images. The black line indicates the standard deviation. (i) Confusion matrix of results with AI-focused images in non-myositis. NA indicates no answer. (j) Confusion matrix of results with AI-unfocused images in non-myositis.

It is difficult to diagnose rare diseases because very few experienced specialists are available; therefore, there is an urgent need for accurate and cost-effective technological diagnostic systems can be used in remote regions^2^. Herein, we report a novel AI-based, CNN-assisted system to pathologically diagnose rare diseases using an algorithm that trained with limited image data (H&E-stained, CCD-shot slides). Our major finding was that the CNNs outperformed human physicians, indicating its potential for cost-effective clinical use, especially in underserved areas.

Since most IIMs are treatable and hereditary muscle diseases are untreatable, accurately differentiating between them is very important, but is often challenging even for experts (e.g., IMNM clinically and pathologically mimics muscular dystrophy, especially in children^19, 20^). Moreover, each IIM subtype requires specific treatment, further highlighting the need for differentiation. In this study, CNNs successfully differentiated IIMs from other muscle diseases and classified IIM into its 4 subtypes (IBM, IMNM, DM, and ASS), suggesting that the system was effective for IIMs. We did not include polymyositis in this analysis because muscle pathologists are increasingly skeptical about its histopathological definitions; this is a recent conceptual change in IIMs^20-22^.

The CNNs also classified 7 major hereditary muscle diseases, demonstrating that the system is compatible with conventional muscle pathology diagnoses. This also suggests that genetic differences can be computationally predicted based on histological features. Further, an accurate pathological diagnosis can help narrow the scope of genetic testing. Hereditary diseases have been diagnosed previously using a combination of pathological and genetic testing^23^.

To visualize the accuracy of the CNNs’ predictions, Coudray *et al*. manually highlighted cropped image patches from a whole-slide photo^5^. In this study, we used Grad-cam^17^ to automatically identify the critical regions for CNN predictions and create images to help investigate the relationship between the CNN predictions and the physicians’ diagnoses.

AI-focused IIM areas were useful for physicians and CNNs; however, there was no significant difference in the non-myositis images. We speculated that the physicians were not as accurate as CNNs for non-myositis diseases because; (1) CNNs may have considered findings that were unknown to the physicians; (2) CNNs may have recognized microstructures, such as rimmed vacuoles and nemaline bodies, that are important diagnostic features of hereditary diseases and are usually recognized by physicians at high magnifications; and (3) physicians are usually trained to observe muscular findings on stains other than H&E. The pathological findings of IIM, e.g., perifascicular atrophy in DM, are large enough to observe even in low-magnification H&E images, but hereditary muscle diseases are easier to identify with other stains, such as the modified Gomori trichrome stain (vacuoles and nemaline bodies) and nicotinamide adenine dinucleotide dehydrogenase (NADH)lJtetrazolium reductase stain (central cores).

In this study, we collected data from one of the world’s largest muscle biopsy collections. However, there are some limitations to it. The performance of CNNs can be influenced by differences in the image data collection, staining protocols, and cameras at various centers. Therefore, it is necessary to conduct further studies with a larger number of pathological images from several laboratories. We expect that, in the future, CNNs will be used in prospective studies and will be embedded directly into medical equipment, as has been done in augmented reality microscopes^24^. We believe that this study provides promising outcomes that support the use of an AI-assisted system for diagnosing neuromuscular disorders.

## Data Availability

Detailed data will be available upon request.

## Acknowledgments

This study was supported by the Japan Agency for Medical Research and Development (AMED) under grant number JP18ek0109348.

## Author contributions

Y. K. designed and conducted the experiments and wrote the manuscript; M. O. reviewed pathological and genetic data and performed the physicians’ test; H. N. and S. Y. analyzed the data and contributed to writing the manuscript; M. I., M. O., Y. S., J. T., A. I., T. K., YL. C., W. Y. and S. H. performed the physicians’ test; T.I. designed the deep learning algorithm; Y. T. prepared the experimental environment; R. T. and A. T. directed the project; F. M. contributed to obtaining grant and designing the framework of the study; I. N. made the pathological diagnoses for all cases and supervised the study.

## ONLINE METHODS

### Dataset

We utilized H&E-stained frozen muscle sections on glass slides that were mounted for diagnostic purposes in the National Center of Neurology and Psychiatry between 1981 and 2019. All materials used in the present study were obtained for diagnostic purposes with written informed consent. The study was approved by the Ethics Committee of the National Center of Neurology and Psychiatry. The samples had already been evaluated by immunostaining, western blot, blood biochemistry, genetic testing, etc.

### Data preparation

Images were taken from slides with CCD cameras (Olympus DP72/DP74) attached to the microscope. The objective lens magnification was 4×, and the image size was 1024 × 1360 or 1600 × 1200. Three pathological images were taken per slide to maximize the coverage of the sample and minimize overlaps between them. If the sample size was small enough to fit into the finder field, only one image was taken.

In contrast, more than three shots were obtained if the sample size was too large. In total, 4,041 images were obtained from 1,400 slides. When the training and validation sets were created, two image patches (1024 × 1024) were cropped from both the left and right edges of the images for data augmentation. Meanwhile, an image patch (1024 × 1024) was cropped from the image center when the test sets were created. The number of images in each shot is presented in **Extended Data Table 1**.

### Deep learning algorithm and architecture design

The detailed architecture of CNN in this study is presented in **Extended Data Figure 1**. The input images were resized from 1024 × 1024 to 640 × 640. The classifier internally divided an input image into sixteen 160 × 160 image patches and created 16 feature maps. Owing to the architecture, the training data for feature generators was multiplied by 16 times. Densely connected convolutional networks^25^, called DenseNet, were used as feature generators in this study.

DenseNet generally consists of dense blocks and transition layers. A dense block has two convolutional layers and a concatenation layer. A transition layer has a convolutional layer and an average pooling layer. Our DenseNet has 58 dense blocks and three transition layers. The DenseNet was pre-trained with ImageNet^26^, an extensive image database. All feature maps were concatenated, averaged by global average pooling, and used to generate the final probability. At the same time, each feature map was used to generate the probability per image patch. In the test phase, only the final probability was used for the prediction. In the training phase, the final probability and probabilities of image patches were used to calculate the loss function (L) as:

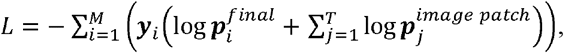

where *M* is the number of classes, *T* is the number of patches, ***y*** is a one-hot vector (true class is 1 and others are 0), ***p*** ^*final*^ is a vector of the probability of final prediction, and *p* ^*patch*^ is a vector of the probability of prediction by image patch.

### Training and testing the algorithm for the IIM differentiation task

For this task, we adopted the holdout method that divided the data into a training and test set. We randomly selected 96 slides as a test set and used the remaining 1,304 slides as a training set. In the training phase, the training set was divided into five groups; four were trained and one group was used to validate the progress of the training set. We carried out the training five times while shifting the evaluation set one by one to train five CNNs (**Figure 1i**). In the test phase, we used a ‘model ensemble’ method^18^ that averaged the probabilities of images cropped from the same slide to obtain the image probability by one slide and also averaged the output probabilities of the five CNNs. The probability averaging function is:

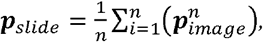

where *n* is the number of images per slide, ***p***^*slide*^ is a vector of the probability of a slide, and ***p***^*image*^ is a vector of the probability of an image. The class with the highest probability was adopted as the final prediction.

The following function of averaging the probabilities of models is:

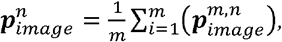

where *m* is the number of models and 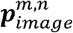 is a vector of the probability of an image outputted by *a* model. Furthermore, ***p***_*slide*_ is:

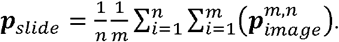

The disease that had the highest probability was selected as the final prediction.

### Setting the training algorithm

The number of trainings (the ‘epoch size’) was 60. Loss and accuracy were calculated using the validation set in every training. The CNN that had the best accuracy was chosen for the test phase. We used rectified Adam^27^ as the optimizer because it is more robust against the variance of learning rates than the Adam^28^ optimizer. The initial learning rate of the optimizer was 0.0001. CNNs were trained on Nvidia V100 using Tensorflow 1.13.2 (https://github.com/tensorflow/tensorflow) and Keras 2.2.4 software (https://github.com/keras-team/keras).

### Transfer learning

Transfer learning^29^ is a technique employed to improve the performance of deep neural networks when training data is limited. It operates by using the parameters of CNNs that are trained in one task and applying them to another task. In this study, the CNNs trained in IIM differentiation were used to classify IIMs and non-myositis muscle diseases; first, the CNNs were trained in IIM differentiation; second, the final output layer of the trained CNNs was changed from two units to four units because there were two classes in the IIM differentiation task and four in the IIM classification task; third, the CNNs were trained in IIM classification. Transfer learning can be implemented in one of two ways: one is by having fixed the parameters and excluding the output layer, and the other is by having unfixed parameters. We used the unfixed approach while classifying IIMs and non-myositis muscle diseases and set the number of output units to 7.

### Metrics

We evaluated the performance of the algorithm using the following metrics: accuracy, ROC curves (true- versus false-positive rate), and AUC (area under the ROC). The accuracy was calculated as follows:

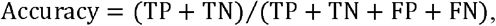

Where TP is true-positive, TN is true-negative, FP is false-positive, and FN is false-negative. When we used the 5-fold cross-validation, we summed all the TP, TN, FP, and FN values in five shots. ROC curves were generated by sweeping the threshold from 0 to 1. In multi-class classifications, one target class was set as a positive class and the other classes were negative. ROCs of IIM and non-myositis muscle disease classifications are presented in **Extended Data Figures 2 and 3 and Extended Data Table 1**.

### Visualization

In previous studies, heatmaps were generated to visualize the prediction by deep CNNs^17, 30, 31^. In this study, we adopted Grad-cam, which can generate visual explanations from any CNN-based network without requiring architectural changes or re-training. This method uses gradients between confidences and feature maps to identify reactive filters. The gradients are calculated as:

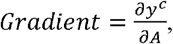

where *y*^*c*^ is the confidence of class *c*, and *A* is a feature map. We used the following function to calculate gradients because the aggregated classifier internally generated feature maps per image patch:

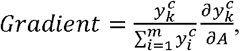

where *m* is the number of image patches, and 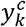 is *y*^*c*^ of image patch *k*. The representation ability of Grad-cam depends on the size of the feature map: the larger the size of a feature map, the better the representation. The size of the feature map in this study was 5 × 5, but the aggregated classifier generates 16 (4 × 4) feature maps, which means that, practically, the size of the feature maps was 20 × 20.

### Visualization test

We created masked images to investigate the relationship between the physicians’ predictions and diagnoses by; first, generating heatmap images with Grad-cam from H&E-stained images; second, calculating the median for each heatmap image; and third, transforming some heatmap images into mask images by masking areas below the median and applying them to H&E-stained images to create AI-focused images, and creating the remaining mask images by using the same method to obtain AI-unfocused images. The number of AI-focused IIM images, AI-unfocused IIM images, AI-focused non-myositis muscle disease images, and AI-unfocused non-myositis muscle disease images was 25, 25, 24, and 24, respectively.

Seven physicians differentiated the IIM images into four subtypes (ASS, IBM, DM, and IMNM) and non-myositis images into seven subtypes (DYST, LG2A, LG2B, CM, FCMD, UCMD, and GNEM). The pathologists were not informed about which images were AI-focused during testing. Each pathologist’s accuracy was calculated and averaged in each group to calculate the significant difference between AI-focused and AI-unfocused images using the paired *t*-test. The function of the test was:

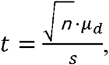

where *µ*_*d*_ and *s* are the mean and standard deviation of differences between all pairs and *n* is the number of samples. We assumed the results of the pathologists’ normally distributed data.

### Participating physicians

Nine physicians (three adult neurologists, four pediatric neurologists, and two pathologists) who were specially trained in muscle pathology differentiated between IIM and other conditions by studying 96 pathology slides. The training period for muscle pathology ranged from 1–23 years (average: 5.5 years). For the visualization test, seven physicians (excluding two adult neurologists from the nine physicians) diagnosed 98 cases by studying AI-focused or AI-unfocused images.

## EXTENDED DATA LEGENDS

**Extended Data Figure 1** | **Detailed design of the deep convolutional neural network**. The architecture is based on an aggregated classifier. Densely connected convolutional networks, called DenseNet, were adopted as a feature extractor. DenseNet generally consists of dense blocks and transition layers. A dense block has two convolutional layers and a concatenation layer, and a transition layer has a convolutional layer and an average pooling layer. The study used DenseNet that had 58 dense blocks and three transition layers.

**Extended Data Figure 2** | **ROC curves of all shots of 5-fold cross-validation of IIM classification**. Left to right in the upper line: IMNM (a), and IBM (b). Left to right in the lower line: ASS (c) and DM (d).

**Extended Data Figure 3** | **ROC curves of all 5-fold cross-validation shots of non-myositis muscle disease classification**. Left to right in the upper line: CM (a), FCMD (b), and GNEM (c). Left to right in the middle line: LG2B (d), UCMD (e), and DYST (f). The lower line: LG2A (g).

**Extended Data Table 1.** Amount of data required for training and validation and the results of every test case. The test data following IIM differentiation was the same because they were fixed to be compared with physicians. The amount of training and validation data were increased by data augmentation. Performance per slide was better than performance per image in most of the test cases.

| Test case | Test type | CV No. | Data (image) |  |  | Per slide |  | Per image |  |
| --- | --- | --- | --- | --- | --- | --- | --- | --- | --- |
|  |  |  | Train | Val | Test | AUC | Accuracy | AUC | Accuracy |
| IIM differentiation | Ensemble | N/A | N/A | N/A | 288 | 0.996 | 0.969 | 0.992 | 0.958 |
|  | 5-CV - Shot1 | 1 | 8364 | 2152 | 288 | 0.988 | 0.948 | 0.987 | 0.941 |
|  | 5-CV - Shot2 | 2 | 8412 | 2104 | 288 | 0.993 | 0.958 | 0.984 | 0.934 |
|  | 5-CV - Shot3 | 3 | 8420 | 2096 | 288 | 0.992 | 0.958 | 0.986 | 0.941 |
|  | 5-CV - Shot4 | 4 | 8408 | 2108 | 288 | 0.995 | 0.948 | 0.990 | 0.958 |
|  | 5-CV - Shot5 | 5 | 8460 | 2056 | 288 | 0.995 | 0.958 | 0.987 | 0.938 |
|  | 5-CV - Average | N/A | N/A | N/A | 288 | 0.993 | 0.954 | 0.987 | 0.942 |
| IIM classification | 5-CV - Shot1 | 1 | 1520 | 500 | 252 | 0.960 | 0.812 | 0.954 | 0.802 |
|  | 5-CV - Shot2 | 2 | 1510 | 504 | 255 | 0.953 | 0.800 | 0.944 | 0.784 |
|  | 5-CV - Shot3 | 3 | 1510 | 510 | 252 | 0.941 | 0.857 | 0.940 | 0.829 |
|  | 5-CV - Shot4 | 4 | 1514 | 504 | 253 | 0.965 | 0.881 | 0.956 | 0.842 |
|  | 5-CV - Shot5 | 5 | 1518 | 506 | 250 | 0.983 | 0.843 | 0.970 | 0.848 |
|  | 5-CV - Average | N/A | N/A | N/A | N/A | 0.958 | 0.839 | 0.970 | 0.821 |
| Classification of non-myositis muscle disease | 5-CV - Shot1 | 1 | 2956 | 944 | 510 | 0.896 | 0.695 | 0.890 | 0.688 |
|  | 5-CV - Shot2 | 2 | 2956 | 1020 | 472 | 0.948 | 0.781 | 0.942 | 0.750 |
|  | 5-CV - Shot3 | 3 | 2992 | 944 | 492 | 0.950 | 0.749 | 0.942 | 0.746 |
|  | 5-CV - Shot4 | 4 | 2908 | 984 | 514 | 0.961 | 0.823 | 0.952 | 0.811 |
|  | 5-CV - Shot5 | 5 | 2948 | 1028 | 472 | 0.920 | 0.676 | 0.913 | 0.669 |
|  | 5-CV - Average | N/A | N/A | N/A | N/A | 0.936 | 0.745 | 0.928 | 0.733 |

