## Supplementary figures and images for "A deep convolutional neural network-based algorithm for muscle biopsy diagnosis outperforms human specialists"

### Extended Data Figure 1

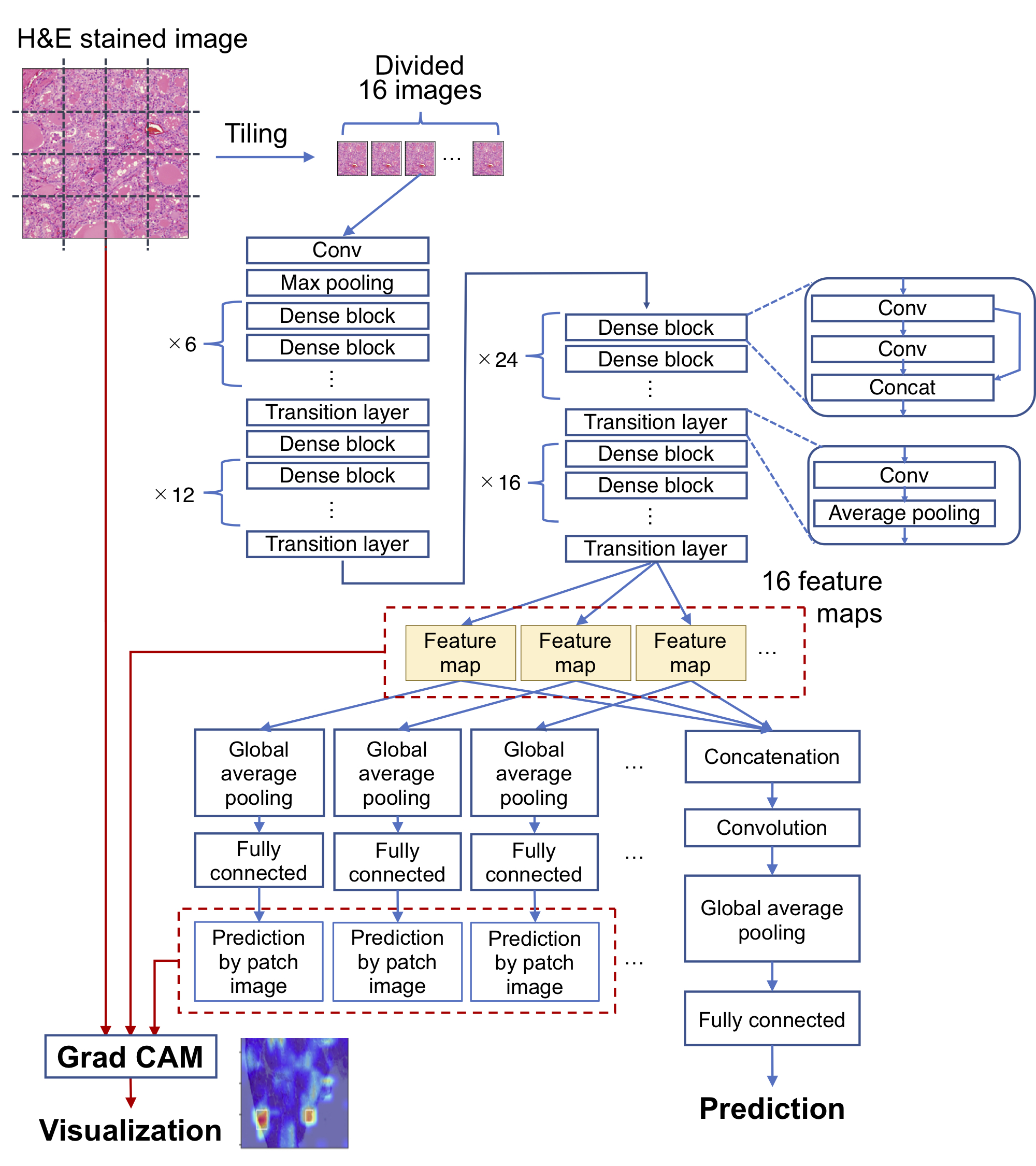

### Extended Data Figure 2

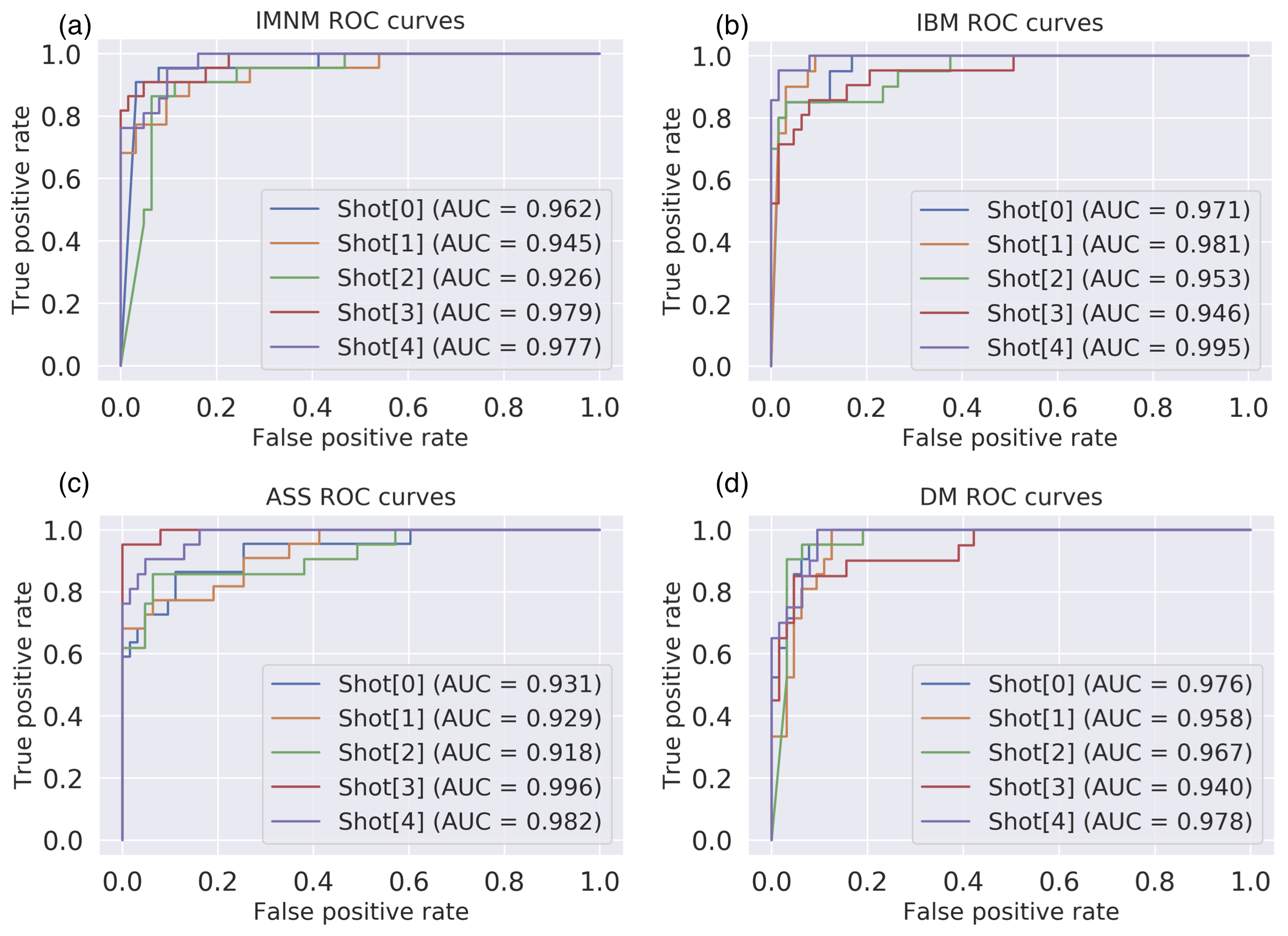

### Extended Data Figure 3

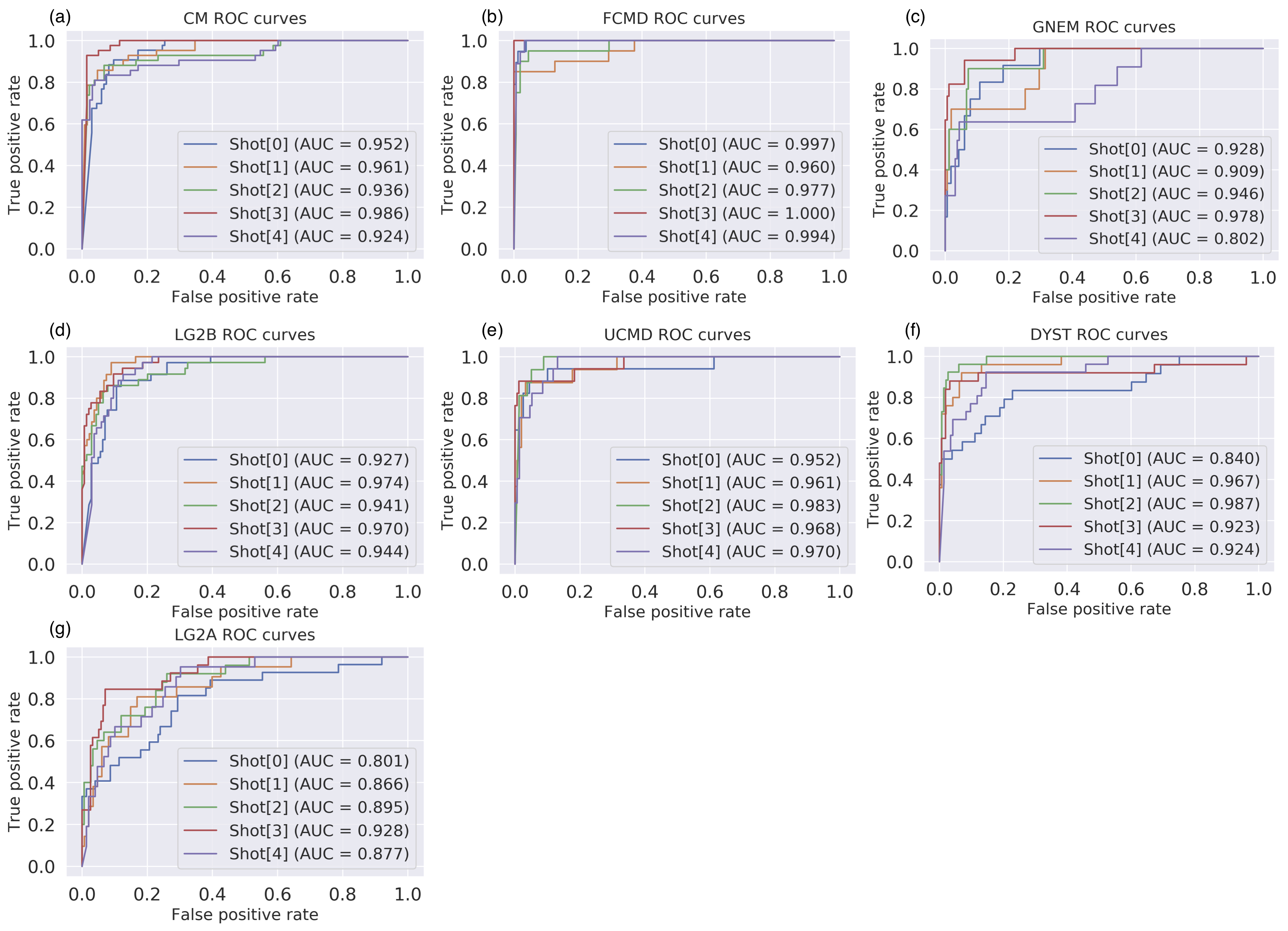
